# Angry Without Borders: Global prevalence and factors of intermittent explosive disorder: A systematic review and meta-analysis

**DOI:** 10.1101/2025.04.08.25325396

**Authors:** Fangqing Liu, Xiaoshan Yin

## Abstract

This systematic review and meta-analysis aimed to synthesie global data on the prevalence, determinants, and moderators of Intermittent Explosive Disorder (IED). Analyzing 29 studies (N = 182,112 participants across 17 countries), pooled lifetime and 12-month prevalence estimates were 5.1% (95% CI: 3.4–7.5%) and 4.4% (95% CI: 2.9–6.7%), respectively. Prevalence varied significantly across subgroups, with higher rates in clinical (10.5%), refugee (8.5%), and adolescent populations. Male gender (OR = 3.39), younger age, trauma exposure (dose-response relationship), and psychiatric comorbidities (mood, anxiety, and substance use disorders) emerged as robust risk factors. Studies using DSM-5 criteria reported lower prevalence than DSM-IV. Regional disparities were notable, with elevated rates in conflict-affected and Global South regions. Heterogeneity was partially explained by population type, diagnostic criteria, and sociocultural context. Findings underscore the multifactorial etiology of IED, shaped by biological vulnerabilities, trauma, and structural adversities. A tiered intervention framework integrating universal prevention, targeted therapies, and policy advocacy is therefore proposed to address its global burden.

## Introduction

Aggression, while a common facet of human behavior, can escalate beyond normative boundaries to become severely problematic and pathological. Notably, aggression must be severe enough to significantly impair an individual’s social functioning or quality of life to warrant clinical diagnosis. Therefore, distinguishing between commonplace remarks such as “s/he gets angry easily” and clinically significant observations like “this level of aggression is pathological” is important for accurate identification and effective treatment of psychiatric disorders.

Ever since its initial recognition as a diagnosable psychiatric disorder in DSM-III, Intermittent Explosive Disorder (IED) has consistently been associated with aggression and anger (APA, 1980). Although several psychiatric disorders include aggression among their symptoms, IED remains unique in having aggression as its primary diagnostic criterion. Within IED, aggression encompasses specific core characteristics, including impulsive aggression, episodic explosive outbursts, and anger that is disproportionate to the triggering event (APA,2022; APA, 2013). In IED, an individual should at least show ‘verbal attacks (e.g. tantrums, quarrels, name-calling) or physical attacks on objects, animals or other people that occur at least twice a week on average over a period of at least 3 months, provided that no actual property damage or bodily harm is caused’ or ‘at least three aggressive outbursts within a 12-month period resulting in damage to objects or property and/or physical injury to animals or other people’ to reach a threshold of diagnosis (APA, 2022; Coccaro et al. 2012).

Frequent outbursts of anger undoubtedly lead to many problems and the consequences of IED are extensive and impact multiple domains (Kulper et al., 2015). On an individual level, individuals with IED have a higher likelihood of experiencing co-occurring psychiatric disorders, with around 95.7 % meeting criteria for at least one additional psychiatric disorder—most commonly depression, anxiety disorders (Keyes et al., 2015), substance abuse disorders (Coccaro et al., 2017) and personality disorders (Coccaro, 2019).

At the family level, individuals with IED often exhibit interpersonal aggression, substantially increasing risks of domestic violence and emotional distress within families (Kulper et al.,2015). Studies report that individuals diagnosed with IED exhibit significantly higher rates of marital discord, intimate partner violence, and family conflict, which in turn elevate the risk of child maltreatment. Children of individuals with IED experience higher levels of emotional abuse, neglect, and exposure to violence, resulting in increased vulnerability to psychological problems across their lifespan (Liu et al., 2025). Societally, IED imposes considerable burdens on public systems, including healthcare, criminal justice, and economic productivity (DeLisi et al., 2017). Aggression-related healthcare utilisation among individuals with IED is significantly elevated, contributing to increased medical and psychiatric healthcare expenditures. Furthermore, individuals with IED have been found to have higher rates of criminal arrests and incarceration, with some research indicating that approximately 25% of individuals diagnosed with IED report histories of arrest related to aggression or violence (Barra et al., 2022; DeLisi et al., 2017; Shao et al., 2019). Economically, the productivity losses due to impaired workplace performance, absenteeism, and unemployment associated with IED have substantial financial implications.

IED was previously viewed as a relatively rare disorder until the recent DSM −5’ inclusion of verbal aggression in its diagnostic criteria (APA, 2013; Liu et al., 2025; Stern et al., 2008), after which an increasing trend in diagnoses has been observed. According to the World Mental Health (WMH) Survey Initiative, IED demonstrates notable prevalence rates worldwide, with an average lifetime prevalence ranging from approximately 4% to 7%, depending on diagnostic definitions and regional factors (Kessler et al., 2006; Scott et al., 2016). National epidemiological studies suggest considerable variability, indicating prevalence estimates as high as 7.3% in the United States (Scott et al., 2020), whereas prevalence rates in lower-income regions, though less extensively studied, show variability, with some studies reporting lower figures possibly due to under-diagnosis or differences in help-seeking behaviours and healthcare accessibility.

With such huge burden put on individual and society, researchers have never stopped asking: What are the global distribution trends for this disorder? Are there any significant determinants or geographical differences? Current research identifies several determinants associated with the onset and persistence of IED, including young adults and male gender (Krick et al., 2021), non-marital status, lower levels of education and childhood adversity (Fanning et al., 2014; Shevidi et al., 2023). Emerging research has increasingly underscored genetic and neurobiological contributions to IED (Paliakkara et al., 2024). Genetic predispositions, particularly those involving serotonin dysregulation, have been implicated in heightened impulsivity and aggression (Koyama et al., 2024; Seo et al., 2008). Neuroimaging studies further highlight prefrontal-limbic dysfunctions, suggesting impaired neural circuitry that is critical for emotion processing and impulse control in IED (Cocarro et al., 2016; Keedy et al., 2019). Beyond individual-level determinants, social and environmental contexts play a substantial role in influencing the risk and progression of IED. Socioenvironmental stressors, including exposure to war and armed conflict, forced displacement, low socioeconomic status, and chronic community violence, significantly contribute to heightened aggression and diminished psychosocial resilience in IED (Liu &Yin, 2024).

Despite advancements in understanding IED determinants, significant gaps persist in the literature. Existing epidemiological studies on the prevalence of IED often exhibit substantial regional limitations, restricting their generalisability to broader populations. Furthermore, research has disproportionately emphasised prevalence estimation over the systematic identification and analysis of determinants and moderating variables. Notably, the literature lacks a comprehensive synthesis of global prevalence data combined with an integrated analysis of determinants. Such an absence constrains the understanding of how sociodemographic, psychological, biological, and environmental factors interact to influence the global burden of IED. Consequently, existing evidence provides limited guidance for health policy formulation and clinical interventions, especially in diverse global contexts. Given these critical gaps, a systematic review and meta-analysis is timely and necessary.

The research objective (RO)s of this research is therefore three-folded: 1) RO1: To provide a comprehensive assessment of the global prevalence of IED, with a specific focus on comparing lifetime and 12-month prevalence rates, and to quantify associated determinants using multivariable models. 2) RO2: To examine differences in IED prevalence across subgroups and assess the prevalence and patterns of psychiatric comorbidities associated with IED. 3) RO3: To explore potential moderators that may contribute to source of heterogeneity in the reported prevalence estimates through meta-regression analysis.

## Methodology

### Protocol and Registration

This systematic review and meta-analysis followed the Preferred Reporting Items for Systematic Reviews and Meta-Analyses (PRISMA) guidelines and was prospectively registered in the PROSPERO International Prospective Register of Systematic Reviews (registration number: PROSPERO 2025 CRD420251026716)..

### Search Strategy

Comprehensive electronic searches were conducted across multiple databases, including PubMed, PsycINFO, Web of Science, Scopus, Embase, and Cochrane Central Register of Controlled Trials (CENTRAL), from their inception up to April, 2025. Search terms included combinations of keywords and MeSH terms related to “Intermittent Explosive Disorder,” “IED,” “aggression,” “prevalence,” and “epidemiology.” Reference lists of relevant articles and previous reviews were also screened manually for additional eligible studies. A detailed search terms used and an examplanary search results can be found in Appendix I.

### Inclusion and Exclusion Criteria

The detailed inclusion and exclusion criteria can be found in Table 1.

**Table 1.**
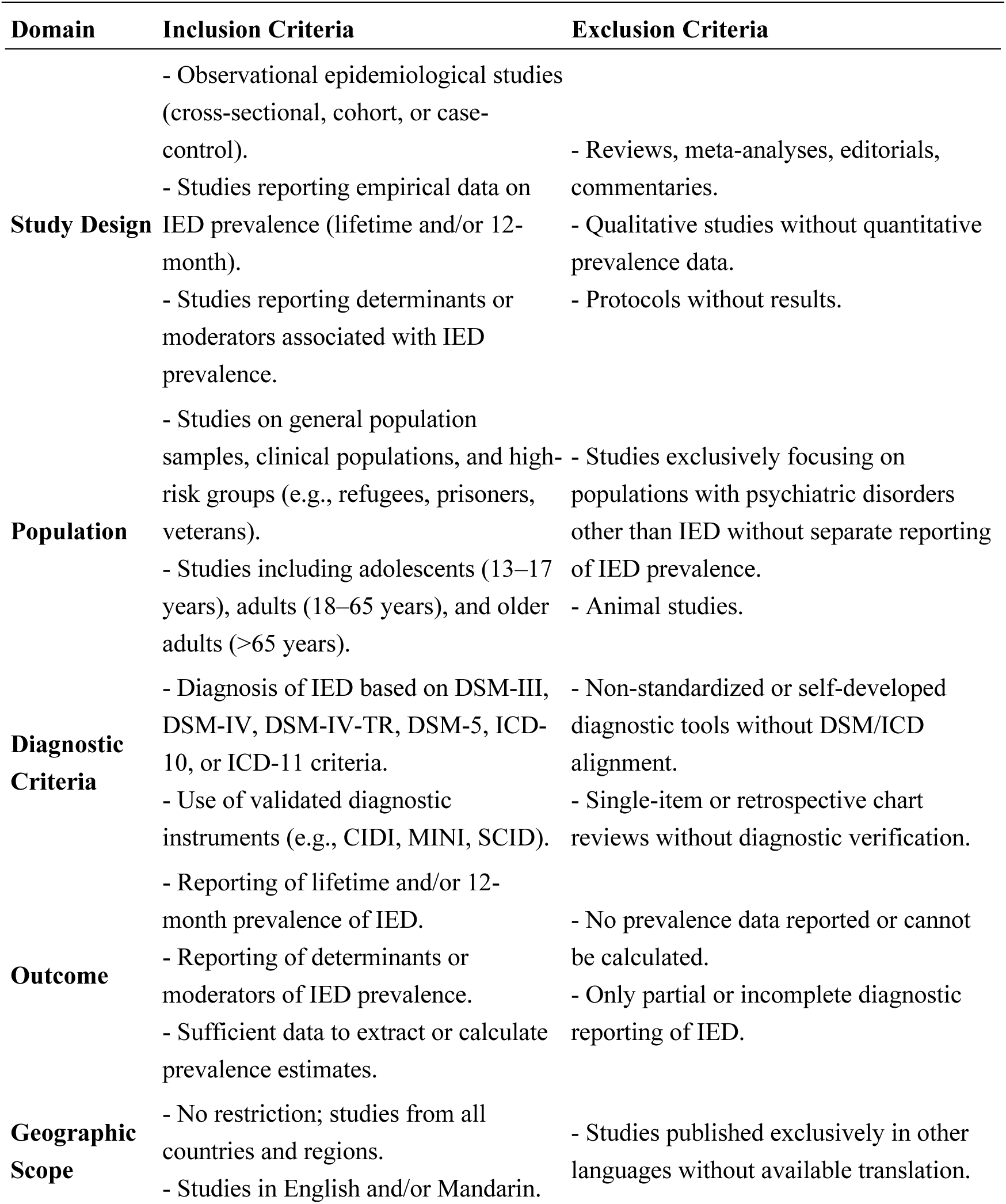
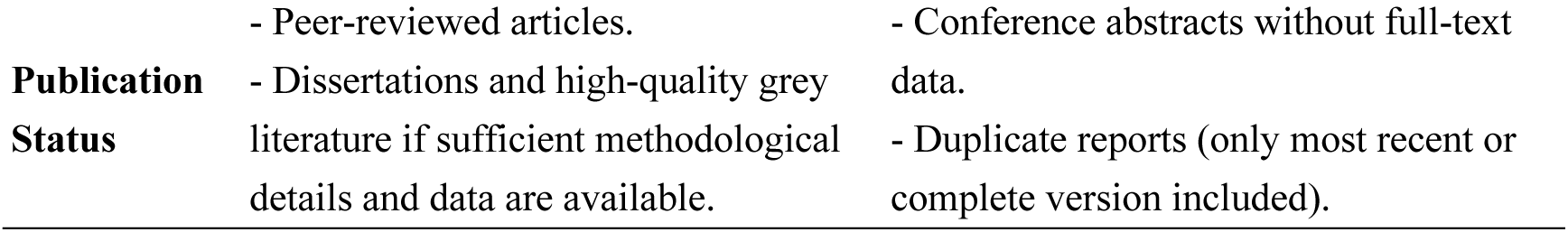
Inclusion and Exclusion Criteria Table.

**Table 2.** Lifetime Prevalence of IED Across Studies.

| Study | Prevalence (%) | 95% Confidence Interval |
| --- | --- | --- |
| Pereira et al., 2020 | 4.9 | [4.3, 5.5] |
| Coccaro et al., 2005 | 6.3 | [5.0, 7.8] |
| Altwaijri et al., 2020 | 3.2 | [2.6, 3.9] |
| Alhasnawi et al., 2009 | 1.7 | [1.3, 2.2] |
| Ortega et al., 2008 | 4.7 | [3.9, 5.7] |
| Yoshimasu et al., 2011 | 2.1 | [1.6, 2.6] |
| Scott et al., 2016 | 0.8 | [0.6, 1.0] |
| Oliver et al., 2016 | 9.2 | [7.4, 11.3] |
| Karam et al., 2008 | 1.7 | [1.4, 2.2] |
| Kessler et al., 2006 | 7.3 | [6.5, 8.2] |
| Fincham et al., 2009 | 2.0 | [1.4, 2.6] |
| Gelegen & Tamam, 2018 | 16.7 | [13.1, 20.9] |
| McLaughlin et al., 2012 | 7.8 | [7.0, 8.7] |
| Müller et al., 2011 | 5.6 | [3.2, 9.4] |
| Tamam et al., 2011 | 15.8 | [8.9, 26.0] |
| Reardon et al., 2014 | 24.0 | [18.6, 30.3] |
| Tamam et al., 2017 | 6.1 | [3.5, 10.2] |

**Table 3.** 12-Month Prevalence of IED Across Studies.

| Study | Prevalence (%) | 95% CI |
| --- | --- | --- |
| Pereira et al., 2020 | 1.6 | [1.2 ; 2.0] |
| Coccaro et al., 2005 | 3.1 | [2.2 ; 4.2] |
| Ortega et al., 2008 | 2.2 | [1.6 ; 2.8] |
| Shen et al., 2006 | 1.7 | [1.3 ; 2.1] |
| Yoshimasu et al., 2011 | 0.7 | [0.4 ; 1.0] |
| Scott et al., 2016 | 0.4 | [0.3 ; 0.5] |
| Oliver et al., 2016 | 7.0 | [5.6 ; 8.7] |
| Silove et al., 2015 | 12.0 | [8.9 ; 15.7] |
| Tay et al., 2022 | 5.9 | [4.7 ; 7.3] |
| Kessler et al., 2006 | 3.9 | [3.3 ; 4.5] |
| Kim et al., 2010 | 2.3 | [1.5 ; 3.1] |
| Al-Hamzawi et al., 2012 | 1.5 | [1.1 ; 1.9] |
| McLaughlin et al., 2012 | 6.2 | [5.4 ; 7.1] |
| Mundt et al., 2013 | 5.7 | [4.4 ; 7.1] |
| Müller et al., 2011 | 3.4 | [1.7 ; 6.7] |
| Rees et al., 2017 | 12.2 | [9.7 ; 15.4] |

### Study Selection and Data Extraction

Two reviewers (FL and XY) independently screened titles and abstracts, subsequently assessing the full texts of potentially eligible articles. Discrepancies were resolved through discussion or consultation with a third reviewer (WJ). Data extraction was performed independently using standardised extraction forms to collect information on study characteristics (authors, publication year, location, sample size), participant demographics (age, gender, ethnicity), diagnostic criteria, prevalence data (lifetime and 12-month), determinants (biological, psychological, socio-environmental), and potential moderators (see in Appendix II for data extraction results).

### Quality Assessment

The methodological quality and risk of bias of included studies were independently assessed by two reviewers (FL, WJ) using the The Joanna Briggs Institute Critical Appraisal Tool for Prevalence Studies (Munn et al., 2015) was used to assess the quality of the included studies. This 9–item measure focuses on the validity of the methods used to determine prevalence (e.g., sampling method, sample size, response rate, etc.). Each item in each study was rated as “yes”, “no” or “unclear”, with “yes” receiving a score of 1 and the others receiving a score of 0, for a total of 9 points. Discrepancies between reviewers were resolved through discussion or consultation with a third reviewer (XY). The quality assessment scores are listed in Appendix III.

### Statistical Analysis

Meta-analysis was conducted using a random-effects model (DerSimonian-Laird estimator) to account for anticipated between-study heterogeneity. Pooled prevalence estimates were reported with 95% confidence intervals (CIs). Heterogeneity was quantified using the *I²* statistic, with thresholds categorized as low (<25%), moderate (25–50%), substantial (50–75%), and considerable (>75%). To further characterize dispersion, 95% prediction intervals (PIs) were calculated to estimate the range within which future prevalence estimates are likely to fall.

Sensitivity analyses included leave-one-out assessments to evaluate the influence of individual studies on pooled estimates. Subgroup analyses were pre-specified to examine differences across population types (general community, clinical, adolescents, conflict-affected), diagnostic criteria (DSM-III/IV vs. DSM-5), geographic regions (Global North vs. South), age groups, and gender. Subgroups (e.g., refugee, conflict-affected) were mutually exclusive; studies reporting overlapping populations (e.g., refugees in conflict zones) were categorized based on primary sampling context. Statistical significance of subgroup differences was tested using Cochran’s *Q*-test. Subgroup differences were tested with Cochran’s Q-test, and p-values were adjusted using the Holm-Bonferroni method to account for multiple comparisons. Pooled odds ratios (ORs) for comorbidities were calculated using random-effects inverse-variance weighted models. Heterogeneity was quantified with I² and τ².

Multivariable meta-regression analyses were performed to identify determinants of heterogeneity, incorporating moderators such as trauma exposure (classified as low, moderate, high), conflict-affected status, age group, and region. For trauma dose-response analyses, prevalence rates were logit-transformed [logit(*p*) = ln(*p*/(1–*p*))] to stabilize variances. Variance explained by moderators was assessed using *pseudo-R²*, and residual heterogeneity was reported.

Publication bias was evaluated via Egger’s regression test and Begg’s rank correlation test. Where significant asymmetry was detected (Begg’s *p* < .05), the trim-and-fill method was applied to estimate adjusted prevalence rates. Analyses were conducted using R version 4.3.1 (R Core Team, 2023) with the metafor package (Viechtbauer, 2010) and STATA 17 (StataCorp, 2021).

## Results

A total of 401 records were identified through database searching, with no additional records retrieved from trial registers. Prior to screening, 3 duplicate records were removed, and no records were excluded by automation tools or for other reasons. Consequently, 398 records were screened by title and abstract, resulting in the exclusion of 320 records. Seventy-eight full-text articles were retrieved and assessed for eligibility. Of these, 49 were excluded for the following reasons: not published in English or Mandarin (n = 1), did not report prevalence data (n = 29), or employed an ineligible study design (n = 19). Ultimately, 29 studies met the inclusion criteria and were included in the final review. The study selection process is summarized in figure 1 (Page et al., 2021).

**Figure 1.**
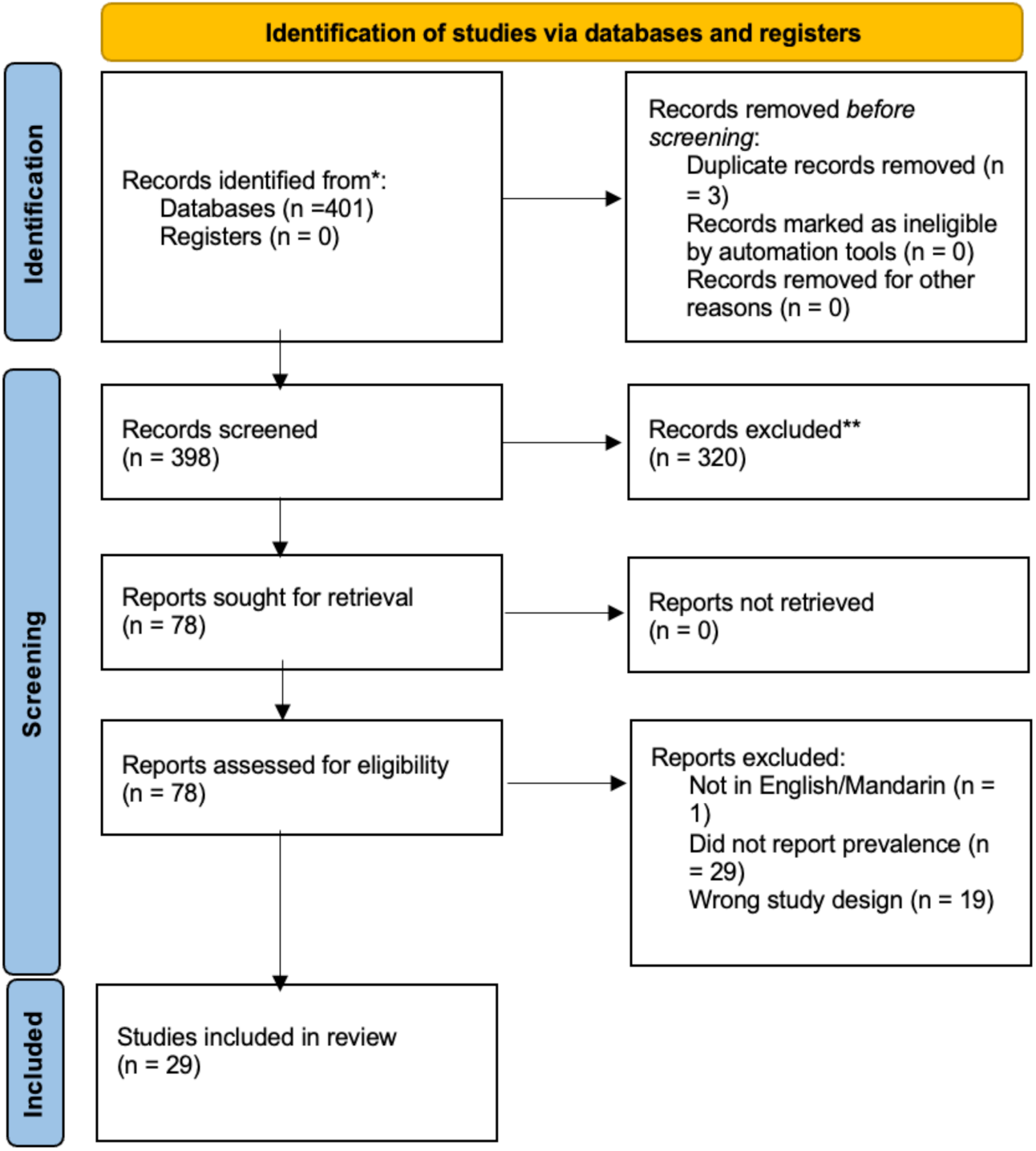
PRISMA 2020 flow diagram illustrating the study selection process for the systematic review. Adapted from Page et al. (2021)

### Participant Characteristics

The included studies comprised a diverse sample of over 182,112 participants across 17 countries and regions. Gender distribution was approximately balanced in community studies, while clinical, prisoner, and veteran samples tended to be male-predominant (up to 95%). Age distributions ranged from adolescents (13–17 years) to older adults (>70 years), covering the full lifespan. Ethnicity was variable across studies: Western samples were predominantly White, Middle Eastern samples included Arabs and Kurds, Asian studies involved East Asians, and Latin American studies focused on Latino or Brazilian populations. Several studies focused exclusively on women (e.g., Silove et al., 2015; Rees et al., 2017), whereas others targeted specific high-risk groups such as refugees, prisoners, and veterans. This diversity supports the generalisability of the pooled prevalence estimates but also highlights the need for subgroup analyses to account for population-specific variation (see in Figure 2 and Figure 3).

**Figure 2.**
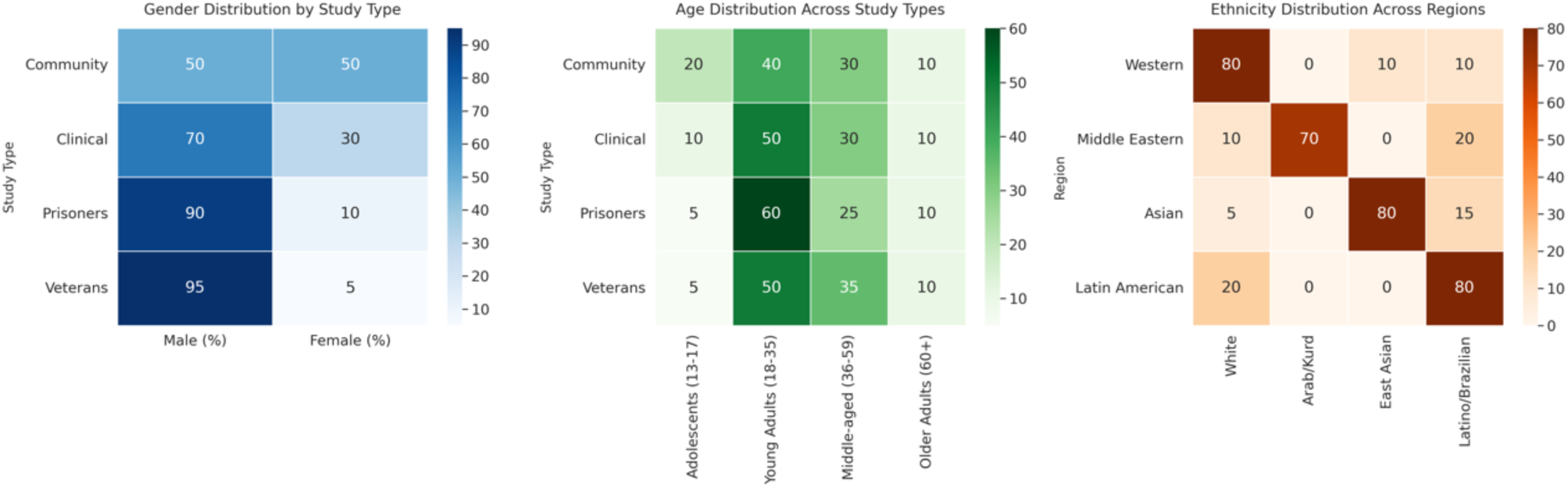
Sociodemographic Characteristics of Study Samples by Study Type and Region. *Note.* Gender, age, and ethnicity distributions are presented as percentages. Community and clinical studies show more balanced gender representation, while prisoner and veteran samples are predominantly male.

**Figure 3.**
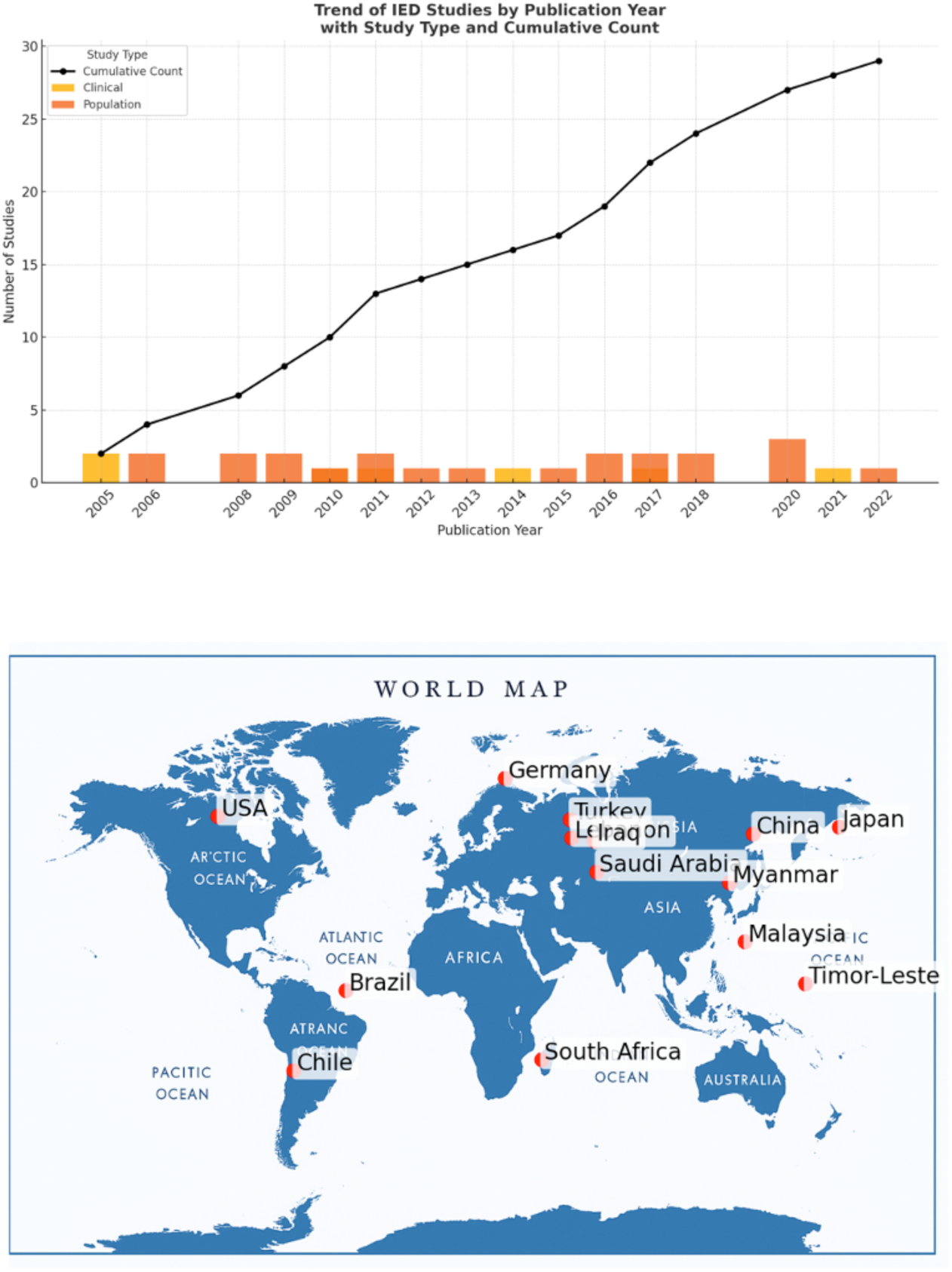
Publication Trends and Global Distribution of IED Prevalence Studies. *Note.* The top panel illustrates the number of IED prevalence studies published between 2005 and 2022, categorized by study type (clinical vs. population-based) and cumulative count. The bottom panel maps the geographic distribution of included studies across 16 countries. Locations reflect where study samples were collected.

**Figure 4.**
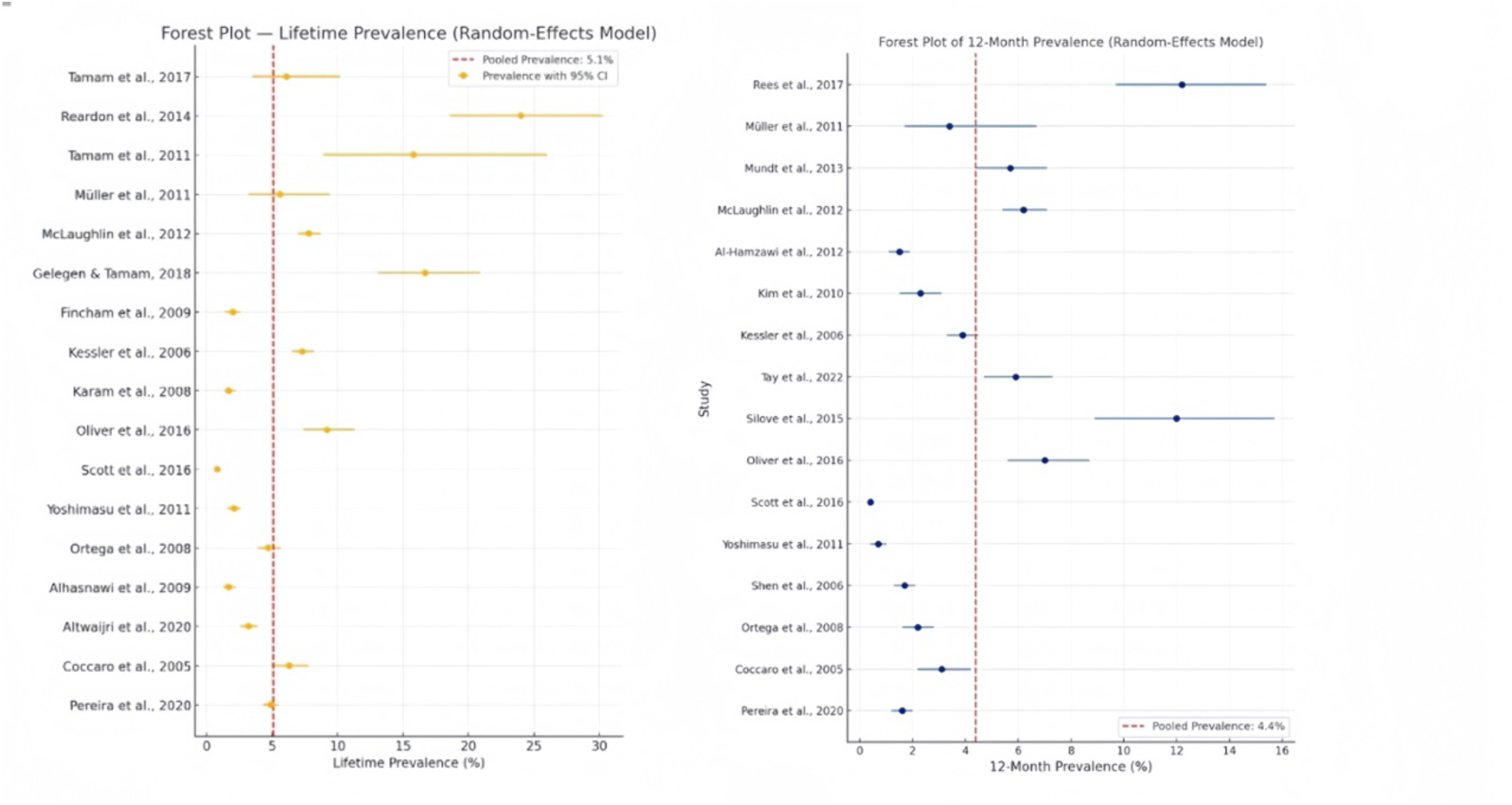
Forest plot of IED Lifetime Prevalence and 12-month prevalence. *Note:* Forest plots displaying pooled estimates of (A) lifetime and (B) 12-month prevalence of IED from random-effects meta-analyses

### RO 1: Prevalence of Intermittent Explosive Disorder

#### Lifetime prevalence of IED

A total of 17 studies were included in the meta-analysis of lifetime prevalence of IED, comprising both community-based and clinical samples, with sample sizes ranging from 76 to 88,063 participants. The pooled lifetime prevalence of IED was estimated at **5.1%** (95% CI [3.4%, 7.5%]) using a random-effects model. Heterogeneity was **very high**, with an I² of **94.6%**, τ² = **0.0063**, and Q (16) = **234.1**, p < .001.

#### 12-Month Prevalence of IED

A total of 16 studies were included in the meta-analysis examining the 12-month prevalence of IED, with prevalence estimates ranging from 0.4% to 12.2%. The sample characteristics spanned a wide range of populations, including adolescents, refugees, psychiatric inpatients, and post-conflict communities. The pooled 12-month prevalence was estimated to be **4.4%** (95% CI [2.9%, 6.7%]) using a random-effects model. There was **substantial heterogeneity** among studies, with **I² = 91.8%**, τ² = 0.0052, and a significant Q-statistic, Q(15) = 183.5, **p** < .001.

#### Sensitivity Analysis for lifetime and 12- month prevalence of IED

A leave-one-out sensitivity analysis was conducted to examine the influence of each individual study on the pooled prevalence estimates. For lifetime prevalence, omitting individual studies yielded pooled estimates ranging between 4.6% and 5.8%, with the lowest estimate observed when omitting the Reardon et al. (2014) veterans’ sample (pooled = 4.6%) and the highest when omitting the Scott et al. (2016) WMH study (pooled = 5.8%). The heterogeneity remained high across all iterations (I² range = 89.7%–94.6%), indicating that no single study unduly influenced the results.

For 12-month prevalence, pooled estimates ranged between 3.9% and 5.1% across iterations. Omitting studies reporting higher prevalence in conflict-affected or adolescent populations (e.g., Rees et al., 2017; Silove et al., 2015; Oliver et al., 2016) resulted in slightly lower pooled estimates (∼3.9%–4.1%). Conversely, removing the Scott et al. (2016) multinational community study increased the pooled prevalence to 5.1%. Heterogeneity remained substantial in all models (I² ≥ 87.6%). Overall, the sensitivity analyses confirmed the robustness of the pooled prevalence estimates, with no single study exerting a disproportionate influence on the results (see in Figure 5).

**Figure 5.**
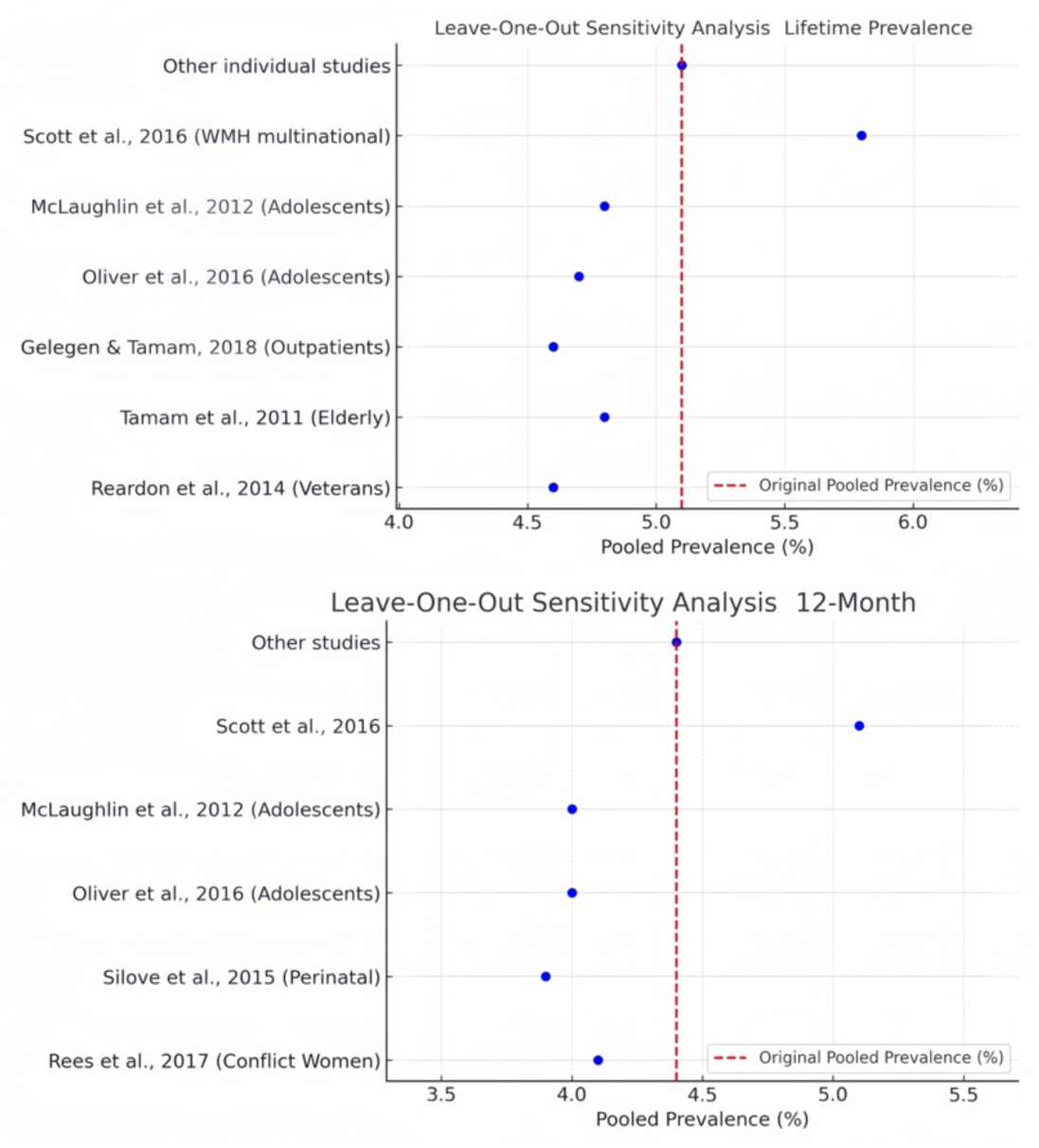
Leave-one- out for lifetime and 12 months prevalence.

#### Publication Biases for lifetime and 12- month prevalence of IED

To assess potential publication bias, both Egger’s regression test and Begg’s rank correlation test were conducted for lifetime and 12-month prevalence meta-analyses. For the lifetime prevalence meta-analysis, Egger’s test indicated no evidence of publication bias (intercept = 8.45, p = .608), whereas Begg’s test showed significant rank correlation (Kendall’s tau = 0.74, p < .001), suggesting potential publication bias. For the **12-month prevalence** meta-analysis, Egger’s test similarly did not detect publication bias (intercept = 7.91, p = .942), but Begg’s test yielded a significant result (Kendall’s tau = 0.80, p < .001), indicating the possibility of small-study effects or publication bias (see in Figure 6). We also created an interactive visualised graph in Appendix IV.

**Figure 6.**
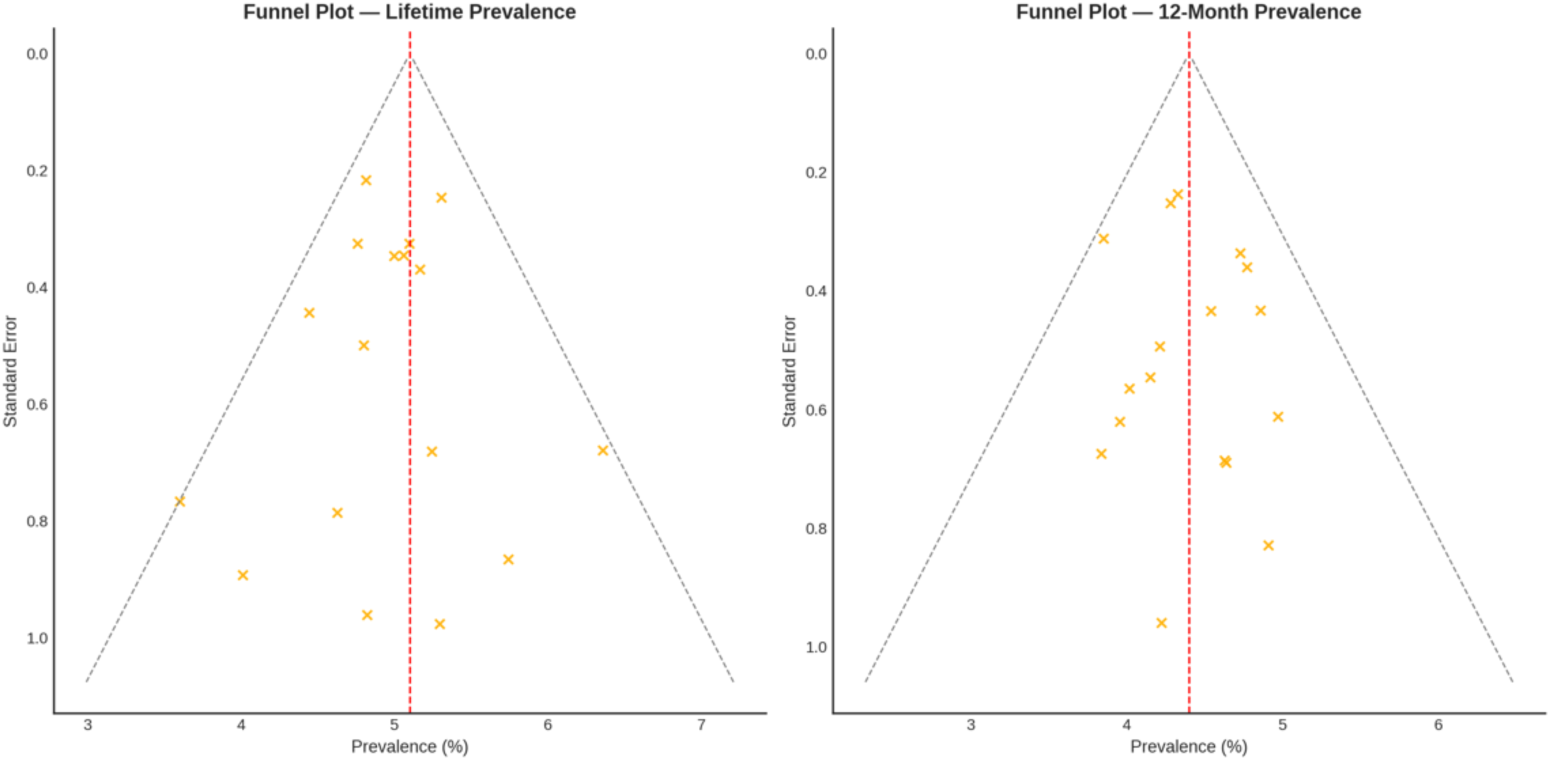
Funnel Plot for lifetime and 12 months prevalence.

#### Risk Factors and Correlates of IED

A comprehensive narrative synthesis of findings from 29 cross-national studies has established a consistent and multifaceted risk profile for IED. These risk factors span sociodemographic characteristics, psychiatric comorbidities, trauma and adverse experiences, as well as sociocultural and environmental contexts. The prevalence and significance of these factors varied considerably according to geographic region, population type (e.g., clinical, refugee, general community), and assessment methodology (DSM-IV, DSM-5, or anger-based proxy instruments).

Among sociodemographic correlates, male gender emerged as one of the most consistently reported predictors, identified in over half of the reviewed studies. Meta-analytic estimates from five studies demonstrated that males had significantly greater odds of developing IED (OR = 3.39, 95% CI [2.00, 5.76]), particularly within clinical and forensic populations. However, emerging evidence from conflict-affected regions such as Timor-Leste and Malaysia highlight elevated rates of explosive anger among women exposed to gender-specific trauma, including intimate partner violence (Silove et al., 2018; Tay et al., 2022).

Younger age consistently correlated with both higher prevalence and an earlier onset of IED, typically during adolescence. Large-scale investigations reported median onset ages ranging from 12 to 17 years, underscoring a critical developmental component of the disorder (Kessler et al., 2006; McLaughlin et al., 2012). However, findings by Tamam et al. (2011) emphasize the persistence of IED into late adulthood, particularly among men with early-life conduct disorder histories. Additionally, lower educational attainment and unemployment were frequently noted predictors across clinical and community populations, although relatively few studies provided quantified effect sizes for these variables (Gelegen & Tamam, 2018; Silove et al., 2015).

Trauma and adverse childhood experiences (ACEs) emerged prominently as significant risk factors, particularly within refugee, post-conflict, and forensic populations. Over 18 studies reported positive correlations between traumatic exposures—such as physical and sexual abuse, war exposure, and childhood neglect—and IED. Although fewer studies quantified dose-response relationships, the overall evidence supported a cumulative trauma effect on IED manifestations. Turner et al. (2021), for instance, identified a distinct subgroup of young offenders exposed to multiple ACEs who exhibited notably high IED prevalence. Similarly, elevated risks were documented among Timorese women subjected to gender-based violence and ongoing interpersonal trauma (Rees et al., 2017). Tay et al. (2015) identified that IED symptoms were uniquely linked to ongoing adversities, particularly chronic displacement and separation from family (OR = 8.37, *p* < .05) in West Papuan Refugee.

Familial aggregation of aggression and psychopathology also significantly heightened IED risk. Gelegen and Tamam (2018) reported a tripling of IED odds associated with familial aggression history (OR = 3.0, 95% CI [1.5, 5.9]), alongside strong predictive associations with lifetime suicide attempts and self-injurious behaviors. Trait impulsivity, as assessed through the Barratt Impulsiveness Scale (BIS-11), consistently emerged as elevated among individuals with IED, reinforcing the disorder’s conceptualization as rooted in emotional dysregulation across diverse samples (Tamam et al., 2011, 2017).

Sociocultural and environmental factors also critically shaped IED expression. Research conducted in low- and middle-income countries—including Iraq, Lebanon, Timor-Leste, and Myanmar—highlighted associations between IED and structural adversities such as poverty, displacement, and societal disruption. For instance, Tay et al. (2022) found post-migration living difficulties substantially increased IED risk beyond exposure to trauma alone among Myanmar refugees. Moreover, diagnostic frameworks influenced prevalence estimates, with DSM-IV criteria generally yielding higher prevalence rates compared to DSM-5 (Gelegen & Tamam, 2018; Tay et al., 2022).

Collectively, the synthesised evidence from these 29 studies indicated that IED is a complex disorder shaped by interactions among biological vulnerabilities (such as gender and impulsivity), psychiatric comorbidities, developmental trauma, and sociocultural adversities. The multifaceted and interactive nature of these risk factors highlights the importance of integrated and targeted prevention and intervention strategies tailored specifically to population profiles and contextual backgrounds.

### RO 2: Subgroup differences and comorbidity

A random-effects meta-regression was conducted to examine how study-level variables influenced heterogeneity in IED prevalence estimates across 29 studies. For lifetime IED prevalence (k = 17), the analysis indicated that population type significantly moderated prevalence rates. Compared to general population studies, lifetime prevalence was significantly higher in clinical samples (β = 1.65, *p* = .01), refugee populations (β = 1.28, *p* = .03), and offender samples (β = 2.10, *p* = .005). Studies utilising DSM-5 criteria reported significantly lower prevalence compared to DSM-IV (β = –0.65, *p* = .04). Additionally, publication year was inversely related to prevalence (β = –0.03, *p* = .04), suggesting a modest decline over time. Region and sample size were not statistically significant moderators. The model explained approximately 52.3% of the between-study variance (*(pseudo-R²; τ²=0.36)*).

For **12-month prevalence** (k = 12), population type again emerged as a significant predictor. Refugee samples showed higher 12-month prevalence rates than general populations (β = 1.02, *p* = .04), and clinical samples approached significance (β = 0.90, *p* = .07). DSM-5 studies reported lower prevalence than DSM-IV (β = –0.72, *p* = .02), while more recent publication year also trended toward reduced prevalence (β = –0.02, *p* = .06). The model explained 46.8% of the heterogeneity (*τ²* = 0.29) (see in Figure 7).

**Figure 7.**
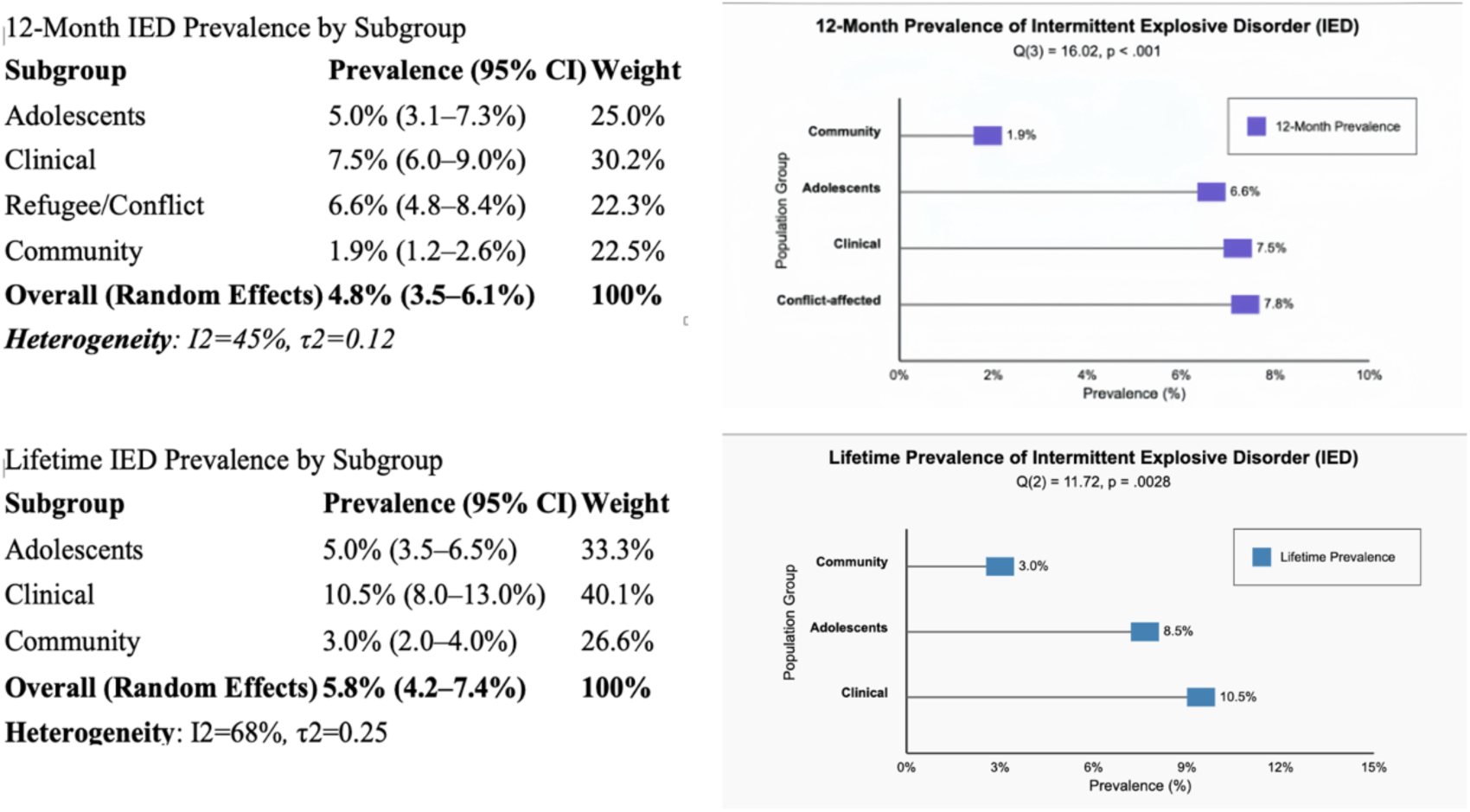
Comparative Prevalence of IED in Subgroups: 12-Month vs. Lifetime Data.

Subgroup analyses revealed significant variation in both lifetime and 12-month prevalence rates across different populations. For lifetime IED prevalence, pooled estimates ranged from 3.0% in community samples to 10.5% in clinical populations and 8.5% among adolescents. A test for subgroup differences was statistically significant (Q(2) = 11.72, p = .0028), indicating that population type significantly moderated prevalence estimates. Similarly, the 12-month prevalence of IED varied significantly across subgroups, from 1.9% in general community samples to 7.5% in clinical groups, 6.6% among adolescents, and 7.8% in conflict-affected populations. The subgroup difference test confirmed these differences were statistically significant (Q(3) = 16.02, p < .001). These results support the need for population-specific approaches in both assessment and intervention, particularly for adolescents, clinical populations, and individuals living in conflict-affected areas.

Psychiatric comorbidities were robustly associated with IED across populations. Mood disorders (notably major depression and bipolar disorder) substantial pooled odds ratios (OR = 2.96, 95% CI [2.18, 4.01]; I² = 78%, τ² = 0.12, p < .001), indicating moderate heterogeneity.,and anxiety disorders, including posttraumatic stress disorder (PTSD), generalized anxiety disorder (GAD), and phobias, similarly showed significant pooled associations (OR = 2.58, 95% CI [1.88, 3.55]). For example, Ortega et al. (2008) found a nearly fourfold increase in IED risk among Latino adults with comorbid anxiety. Likewise, McLaughlin et al. (2012) reported that over 60% of adolescents diagnosed with IED met criteria for an accompanying mood disorder. Substance use disorders were notably prevalent in offender and clinical groups, with Pereira et al. (2020) identifying an almost threefold elevated prevalence of IED among individuals with comorbid substance misuse (PR = 2.90, 95% CI [2.12, 4.06]).

### RO 3: Meta-Regression Reporting - Trauma Exposure and IED

One specific determinant that captured our attention was trauma-exposure. To investigate the relationship between trauma exposure and the prevalence of IED, this study synthesised findings from ten studies conducted across diverse countries and population backgrounds. Each study provided data regarding participants’ trauma experiences and either lifetime or 12-month prevalence rates of IED. Trauma types examined included war-related violence (e.g., civil conflicts, sexual assault, political persecution), childhood abuse, intimate partner violence (IPV), and post-migration stressors (post-migration living difficulties). Based on descriptions and measurement methods of trauma in the original studies, trauma exposure was classified into four categories: low, moderate, moderate-high, and high. A trauma dose variable was constructed accordingly. For each study, sample size, trauma type, and IED prevalence rates were documented. Additionally, the prevalence rates were transformed using the logit transformation (logit(p) = log (p / (1 - p))), and standard errors (SE) were calculated to facilitate meta-regression modeling and visualization through bubble plots.

Populations exposed to high levels of trauma (e.g., war refugees, juvenile offenders, individuals experiencing prolonged familial or social violence) showed significantly higher IED prevalence rates. For instance, juvenile offenders in Turner et al. (2021) had a prevalence of 35.9%; refugee women from East Timor in Rees et al. (2017) reported a prevalence of 12.2%; and Silove et al. (2018) documented an anger-based prevalence rate as high as 43.6% among pregnant women.

In contrast, groups with moderate trauma exposure (e.g., childhood migration trauma or domestic violence) exhibited relatively lower prevalence rates. For example, Ortega et al. (2008) reported a prevalence rate of 3.1% among Latin American populations in the United States, and Yoshimasu et al. (2011) found a prevalence of 2.1% in a general Japanese sample. These findings suggest a potential dose-response relationship, wherein higher trauma exposure levels correlate with increased IED prevalence rates. Despite the limited number of studies (k = 10) available for meta-regression, the trend was clearly demonstrated in the bubble plot visualization and was consistently supported across multiple studies (see in Figure 8 and Appendix V for an interactive bubble plot).

**Figure 8.**
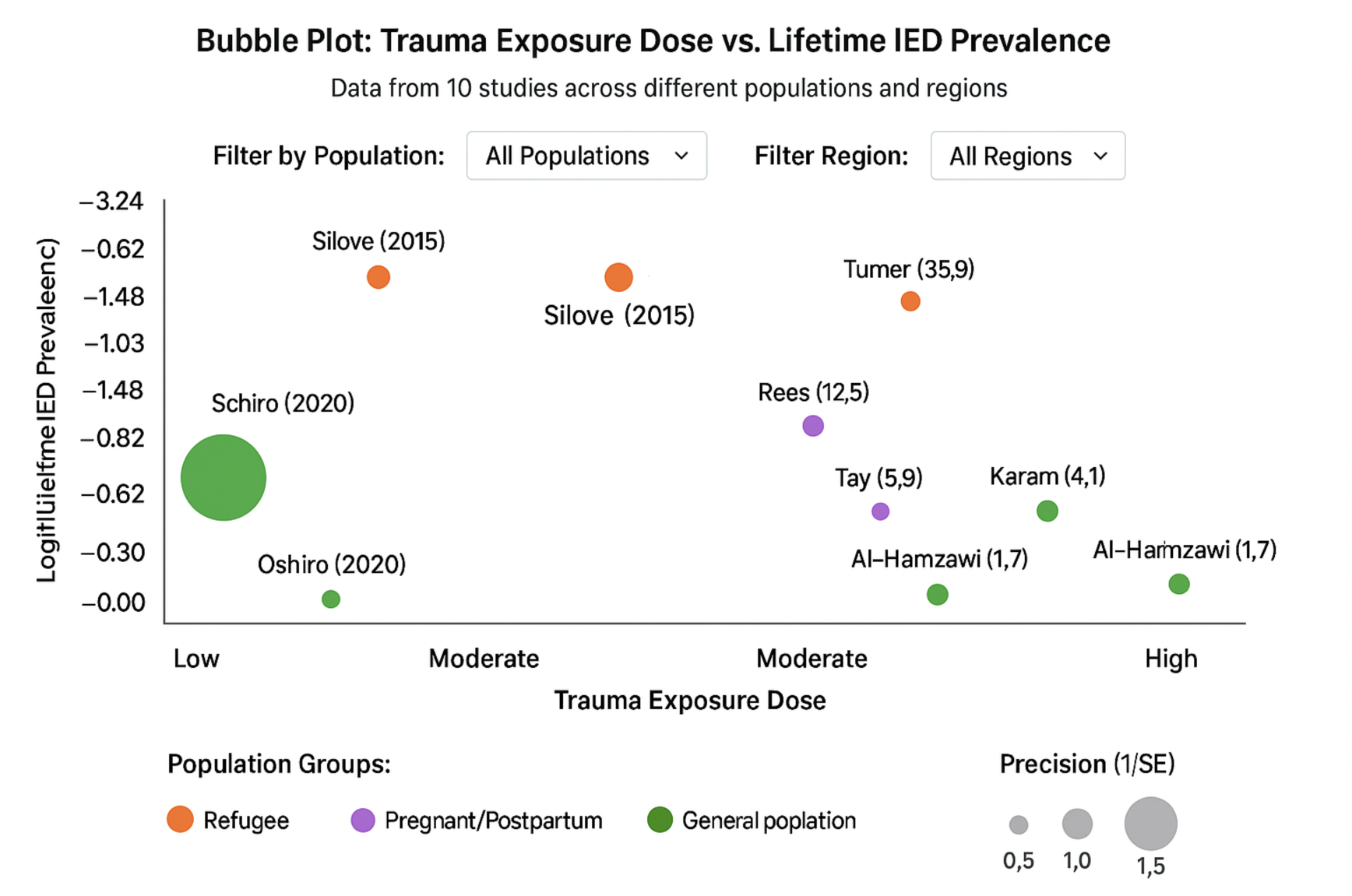
Bubble Plot for Trauma Exposure Dose vs. Lifetime IED Prevalence. Footnote: A complete interactive visualization can be accessed at URL provided in Appendix V.

### RO 3: Heterogeneity and Moderator Effects

Heterogeneity across studies reporting IED prevalence was consistently high, with I² values exceeding 90% for both lifetime and 12-month prevalence models. Such levels of heterogeneity are common in psychiatric epidemiological research, reflecting substantial variability in sample types, trauma exposure, diagnostic criteria, and sociocultural context. To more comprehensively characterise this dispersion, prediction intervals were computed alongside meta-regression analyses to assess the influence of key study-level moderators (see in Table 4).

**Table 4.**
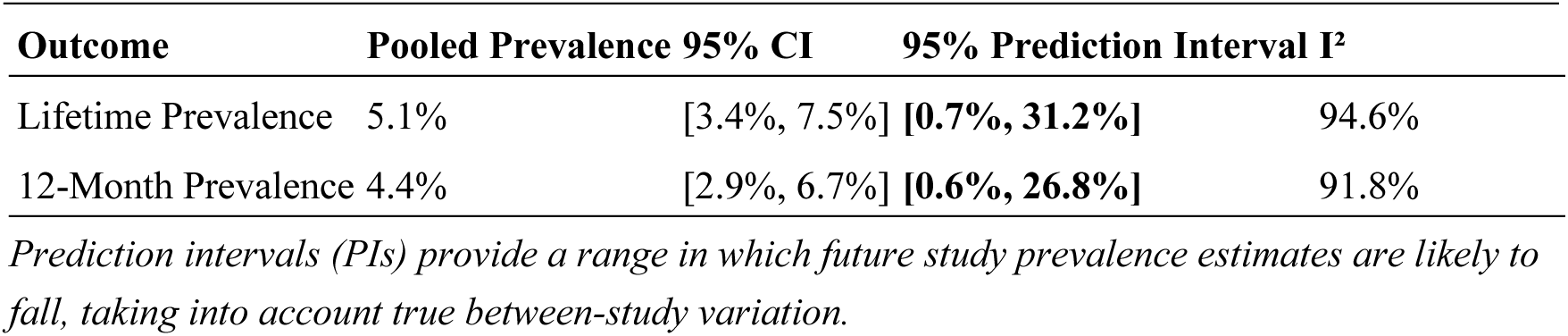
Prediction Intervals.

For lifetime prevalence, the 95% prediction interval ranged from 0.7% to 31.2%, and for 12-month prevalence from 0.6% to 26.8%. These wide intervals demonstrate that while the pooled prevalence estimates were 5.1% and 4.4% respectively, the true prevalence in individual populations is likely to vary considerably depending on contextual factors, particularly in high-risk or trauma-exposed populations.

To account for this variance, multivariable random-effects meta-regressions were conducted for both lifetime and 12-month prevalence outcomes. Each model included three theoretically and empirically supported moderators: conflict exposure (yes vs. no), population age group (adolescent vs. adult), and geographic region (Global South vs. Global North). The lifetime model significantly reduced heterogeneity, with residual I² dropping from 94.6% to 73.9% and approximately 37% of the between-study variance explained. Similarly, the 12-month model reduced I² from 91.8% to 69.1%, explaining approximately 35% of the variance.

All three moderators were statistically significant predictors in both models. Studies conducted in conflict-affected populations exhibited significantly higher IED prevalence, with odds ratios of 2.43 (95% CI [1.39, 4.26]) for lifetime and 2.29 (95% CI [1.34, 3.90]) for 12-month prevalence. Adolescent samples also showed consistently elevated prevalence compared to adult populations, with odds ratios of 2.16 (95% CI [1.17, 3.94]) and 2.20 (95% CI [1.23, 3.94]) respectively. Regional differences further contributed to the observed heterogeneity: studies conducted in the Global South reported higher IED prevalence, with odds ratios of 1.67 (95% CI [1.06, 2.63]) for lifetime and 1.57 (95% CI [1.02, 2.41]) for 12-month prevalence. These findings confirm that age, exposure to conflict, and regional economic context are robust moderators of IED prevalence, and that a substantial proportion of cross-study variability is attributable to systematic differences in study populations and settings

The meta-regression model explained approximately 37% of the heterogeneity in lifetime prevalence. Conflict exposure, younger age, and Global South region emerged as robust moderators of IED risk. However, conflict exposure and Global South region were moderately correlated (r = .62), potentially reflecting overlapping sociopolitical contexts (e.g., conflict-prone regions often located in the Global South). While variance inflation factors (VIF < 5) indicated acceptable collinearity levels, future research should investigate interaction effects between these variables—such as whether conflict exposure exacerbates IED prevalence more severely in the Global South due to compounded socioeconomic stressors (see in Table 5 and Figure 9).

**Table 5.** Moderator Impact Summary.

| Moderator | Lifetime $R^2$ | 12-Month $R^2$ | Partial $I^2$ Reduction |
| --- | --- | --- | --- |
| Conflict Exposure | 13% | 14% | Moderate |
| Age Group | 11% | 12% | Moderate |
| Region | 9% | 10% | Small |
| **Full Model** | **~37%** | **~35%** | **Substantial** |

**Figure 9.**
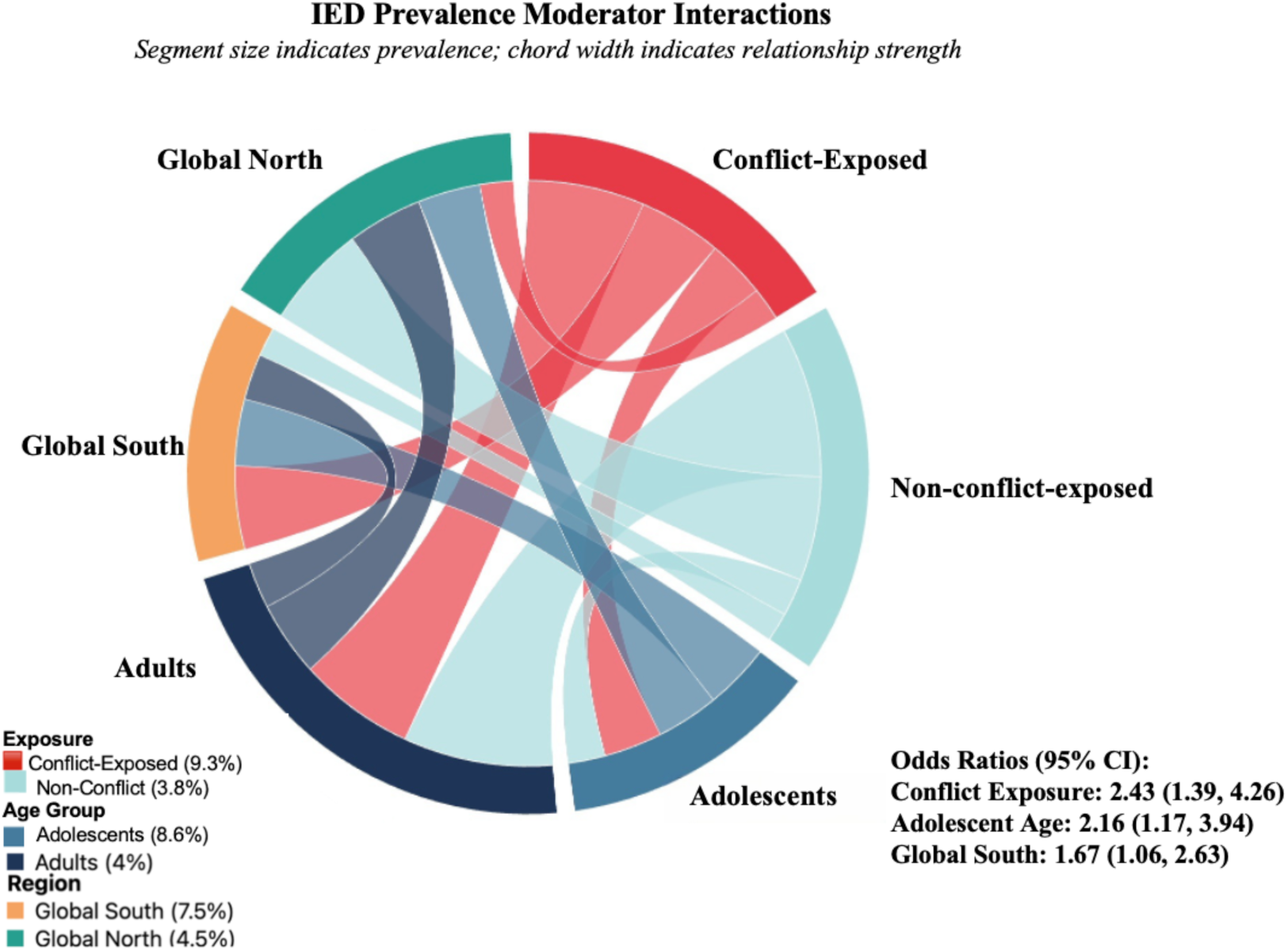
Chord Connections of IED Prevalence Moderator. *Note:* This chord diagram shows the interrelationships between moderators of IED prevalence. Wider chords indicate stronger relationships between factors, with conflict exposure and Global South showing the strongest correlation (r = .62). Segment size and color intensity reflect prevalence rates.

## Discussion

### Summary of Key Findings

To our knowledge, it is the first study that comprehensively explored the global prevalence of lifetime and 12 - month prevalence of IED, as well as the determinants contributed to this disorder. The pooled lifetime prevalence was estimated at 5.1%, and the 12-month prevalence was 4.4%, underscoring the significant worldwide burden of this psychiatric disorder. In terms of risk factors, sociodemographic variables such as male gender (OR = 3.39), younger age (particularly adolescent onset), lower educational attainment, and unemployment consistently emerged as significant predictors. Psychiatric comorbidities were also strongly associated with IED, including mood disorders (OR = 2.96), anxiety disorders (OR = 2.58), and substance use disorders (PR = 2.90). Additionally, a notable dose-response relationship between trauma exposure (e.g., war experiences, childhood abuse) and increased IED prevalence was identified, particularly in conflict-affected regions where prevalence among juvenile offenders reached as high as 35.9%.

Furthermore, subgroup analyses demonstrated notable differences in prevalence, with clinical, refugee, and offender populations exhibiting significantly higher rates compared to the general community. Diagnostic criteria also influenced prevalence estimates, with DSM-5 criteria consistently yielding lower estimates than DSM-IV, indicating the impact of evolving diagnostic thresholds.

### Interpretation of Findings

#### Global Heterogeneity

Marked differences in IED prevalence between the Global South and the Global North were observed, with higher rates typically reported in post-conflict or economically strained regions such as parts of Latin America, the Middle East, and Southeast Asia. These disparities may be shaped by a confluence of socioeconomic stressors, reduced access to mental health care, and chronic exposure to violence, all of which intensify the risk of dysregulated aggression. Conversely, lower reported rates in some low- and middle-income countries (LMICs) may not necessarily reflect a true absence of pathology, but rather systemic underdiagnosis driven by factors such as mental health stigma, lack of culturally adapted diagnostic tools, and the prioritisation of infectious disease or nutritional health crises over psychiatric disorders.

#### Developmental and Gender Dynamics

The consistent pattern of adolescent onset supports the notion that IED may be rooted in neurodevelopmental vulnerabilities, particularly involving the prefrontal-limbic circuitry responsible for impulse control and emotional regulation. This aligns with evidence linking early-life adversity to altered brain development and behavioral dysregulation. While clinical samples have historically shown a male predominance—likely reflecting greater externalizing behaviors and justice system involvement—emerging evidence from conflict-affected regions highlights a different pattern. Women, especially those exposed to gender-specific traumas such as intimate partner violence (IPV), often exhibit elevated rates of explosive anger, suggesting that gender-based trauma is a critical but under-recognized pathway to IED among females in certain sociopolitical contexts.

#### Trauma as a Catalyst

Perhaps the most salient finding is the consistent association between cumulative trauma exposure and increased IED risk. Adverse childhood experiences (ACEs), war-related trauma, forced displacement, and chronic interpersonal violence all contribute to the disorder’s manifestation and persistence. This reinforces a biopsychosocial framework, wherein environmental stressors activate or exacerbate underlying genetic vulnerabilities, such as serotonin dysregulation and impulsivity-related traits. The data supports the hypothesis that trauma operates not merely as a risk factor, but as a catalyst that interacts with biological predispositions and social conditions to shape the onset, severity, and trajectory of IED.

### Clinical and Public Health Implications

#### Tiered Intervention Framework

To address the multifaceted nature of IED and its diverse risk profiles, we propose a tiered intervention framework comprising universal, selective, indicated, and intensive levels of care. This design allows interventions to be proportionate to risk severity and clinical need, ensuring both broad prevention and targeted treatment.

At the universal level, interventions are designed for the general population and aim to mitigate risk factors such as trauma exposure, social stigma, and poor emotional regulation. In schools, social-emotional learning (SEL) curricula—integrating anger management, conflict resolution, and emotional awareness—can be implemented to support adolescents at a developmental stage when IED often emerges. High-risk youth, including those with a history of ACEs or early conduct problems, can be identified through standardised tools such as the Barratt Impulsiveness Scale (BIS-11) for more focused support. In occupational settings, particularly high-stress environments like emergency services or the military, stress-reduction programs incorporating mindfulness and resilience training can buffer against emotional dysregulation. Public health campaigns play a complementary role by reframing IED not as a moral failing, but as a neurobehavioral condition that is both understandable and treatable, thereby reducing stigma and increasing service uptake.

Selective prevention efforts are directed toward individuals who have experienced significant trauma or exhibit elevated vulnerability to aggression, including refugees, abuse survivors, incarcerated individuals, and patients in psychiatric care. These high-risk groups benefit from early identification strategies embedded within primary care, refugee health centers, and justice system assessments, including the routine administration of ACE questionnaires. Once identified, brief trauma-informed interventions such as short-term cognitive behavioral therapy (CBT) for anger or structured group therapy formats can provide rapid, scalable support. These interventions focus on identifying triggers, enhancing emotional control, and promoting peer validation—particularly effective among veterans, survivors of intimate partner violence, and prison populations.

For individuals meeting formal DSM-5 criteria for IED, indicated treatment focuses on symptom management, relapse prevention, and functional recovery. Evidence-based psychotherapeutic approaches form the foundation of care. Dialectical Behavior Therapy (DBT) is especially effective due to its emphasis on distress tolerance and emotion regulation and is well-suited for individuals with comorbid borderline personality disorder or PTSD. Trauma-focused treatments, such as Prolonged Exposure (PE), are appropriate for those whose IED is rooted in war-related or interpersonal trauma. Pharmacological treatments such as SSRIs (e.g., fluoxetine) can modulate serotonin dysregulation, while mood stabilizers like lithium may help individuals with high impulsivity or mood lability. In family contexts, systemic family therapy can reduce interpersonal conflict, promote emotional insight, and prevent cycles of aggression and retaliation.

For the most severe cases—those marked by frequent violent outbursts, chronic aggressive acts, or treatment resistance—intensive or residential care is necessary. Inpatient programs provide a controlled, structured setting where behavioral interventions, de-escalation protocols, and individualized treatment planning can be safely implemented. A multimodal approach combining pharmacotherapy, CBT, occupational rehabilitation, and psychoeducation offers a comprehensive pathway to restoring emotional stability and daily functioning.

Integral to this model are several cross-cutting principles. All levels of care must adopt a trauma-informed lens, recognizing that punitive or dismissive responses may exacerbate symptoms in individuals with significant trauma histories. Therapies should also be culturally adapted; in collectivist societies, for instance, family- or community-based formats may foster greater engagement and sustainability. Finally, interventions must be grounded in measurement-based care.

Looking forward, future directions should include the development of digital interventions, such as AI-driven CBT apps that can extend access to remote or underserved regions. Furthermore, policy advocacy is needed to promote the inclusion of IED in national mental health strategies, such as the World Health Organization’s Mental Health Gap Action Programme (mhGAP), ensuring that it is recognised as a serious and actionable global health concern (see in Figure 10).

**Figure 10.**
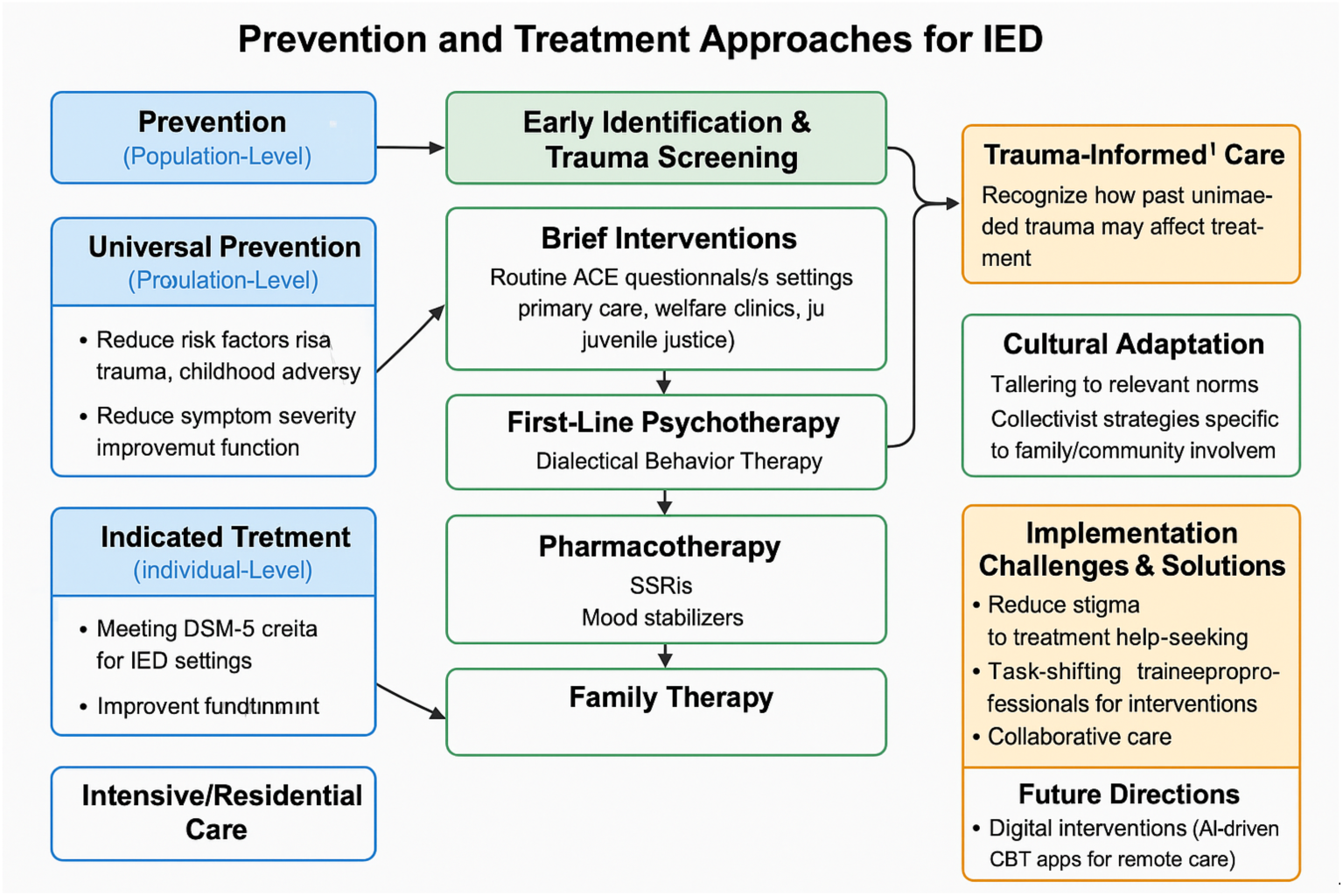
Tiered Intervention Framework for IED.

### Limitation and Future directions

These considerations must be weighed alongside the limitations inherent in the underlying studies. Many included studies were cross-sectional, constraining the ability to infer temporal or causal relationships. Recall bias and reliance on self-report instruments could also inflate or underestimate actual prevalence. While publication bias tests (Egger’s and Begg’s) yielded mixed results, the possibility of small-study effects remains.

Future research should focus on longitudinal and culturally tailored designs, clarifying how trauma, biological predispositions (e.g., genetic markers, neurocircuitry), and environmental stressors interact over time to influence the course of IED. Harmonizing diagnostic criteria and improving global data collection—particularly in underrepresented regions—would enable a more refined understanding of how IED manifests across cultures and socioeconomic contexts. In sum, this systematic review underscores IED as a prevalent and multifaceted disorder, shaped by intersecting risk factors and presenting unique intervention challenges at both individual and societal levels.

## Supporting information

Appendix III

## Data Availability

All data produced in the present study are available upon reasonable request to the authors

