## Appendix I for "Angry Without Borders: Global prevalence and factors of intermittent explosive disorder: A systematic review and meta-analysis"

Appendix I-Exemplary Search

**Exemplary Search Results**

APA PsycInfo <1806 to March 2025 Week 4>

EBM Reviews - Cochrane Central Register of Controlled Trials <February 2025>

Ovid MEDLINE(R) ALL <1946 to April 01, 2025>

1 (Intermittent Explosive Disorder or IED or Explosive anger).mp. [mp=ti, ab, hw, kw, tc, id, ot, tm, mf, fx, sh, bt, nm, kf, ox, px, rx, an, ui, sy, ux, mx] 2443

2 (Prevalence or Epidemiology or Incidence or Global prevalence or Worldwide or International or Cross-national).mp. [mp=ti, ab, hw, kw, tc, id, ot, tm, mf, fx, sh, bt, nm, kf, ox, px, rx, an, ui, sy, ux, mx] 4910855

3 1 and 2 260

4 remove duplicates from 3 257

**Exemplary Search Terms**

| **IED Terms** | **Prevalence Terms** |
|---|---|
| Intermittent Explosive Disorder | prevalence |
| IED | lifetime prevalence |
| | 12-month prevalence |
| explosive anger disorder | epidemiology |
| impulsive aggression disorder | incidence |
| aggressive outbursts | rate |
| | frequency |
| | magnitude |
