## Appendix II for "Angry Without Borders: Global prevalence and factors of intermittent explosive disorder: A systematic review and meta-analysis"

Appendix II-Data Extraction Table 29 studies

| Study | Country | Sample Size | Study Design | Diagnostic Tool | IED Prevalence (Lifetime) | IED Prevalence (12-Month) | Key Findings / Notes |
|---|---|---|---|---|---|---|---|
| Pereira et al. (2020) | Brazil (São Paulo) | 5,037 | Population-based survey (WMH) | WHO-CIDI 3.0 | 4.9% (hierarchical) / 5.3% (non-hierarchical) | Reported in earlier paper, estimated around 1.6% | High comorbidity (76.8%); earlier onset than most disorders; strong association with mood, anxiety, and impulse-control disorders |
| Coccaro et al. (2005) | USA (Rhode Island) | 1,300 | Outpatient psychiatric sample | SCID-I/P (DSM-IV) | 6.3% | 3.1% | Early onset (teen years); higher prevalence in males; strong link to lifetime substance use disorder; 80% showed treatment interest |
| Altwaijri et al. (2020) | Saudi Arabia | 4,004 | National mental health survey (WMH) | WHO-CIDI 3.0 | 3.2% (reported in full paper, usual for disruptive disorders class) | Not reported separately for 12-month | Disruptive behavior disorders (including IED) were 11.2% in total; high association with anxiety and mood disorders; early onset common |
| Alhasnawi et al. (2009) | Iraq | 4,332 | National mental health survey (WMH) | WHO-CIDI 3.0 | 1.7% | Not specified | Lowest prevalence among studies; performed under post-conflict conditions; IED part of impulse-control disorders class; high underdiagnosis suspected |

| | | | | | | | |
|---|---|---|---|---|---|---|---|
| Ortega et al. (2008) | USA (Latino population) | 2,554 | WMH-based population survey | WHO-CIDI | 4.7% | 2.2% | Comorbidity high with mood and anxiety disorders; higher prevalence in US-born Latinos |
| Shen et al. (2006) | China (Beijing & Shanghai) | 5,201 | WMH-based population survey | WHO-CIDI (Chinese version) | Not reported | 1.7% | One of the few studies focused on urban China; lifetime not directly reported but 12-month given |
| Yoshimasu et al. (2011) | Japan | 4,134 | WMH-Japan | WHO-CIDI | 2.1% | 0.7% | Narrow definition yielded lower estimates; young age and male gender were risk factors; strong comorbidity with mood and anxiety disorders |
| Scott et al. (2016) | 16 countries (WMH surveys) | 88,063 | Cross-national WMH survey | WHO-CIDI (narrow + impairment) | 0.8% (range 0.1%-2.7%) | 0.4% | Largest cross-national study to date; male gender, young age, unemployment, low education, and exposure to violence were risk factors. High comorbidity (82%); early onset (~17 years); severe functional impact in ~40% of 12-month cases |
| Scott et al. (2020) | Same dataset (subset n=45,266) | 45,266 | Subtype analysis (WMH dataset) | WHO-CIDI (narrow + impairment) | 0.8% | Not specified | Focus on subtypes: 73% involved harm to people. Externalizing disorders (conduct, substance, ADHD) highly comorbid. Suicidality and impairment significantly higher among more violent subtypes |

Appendix II-Data Extraction Table 29 studies

| | | | | | | | |
|---|---|---|---|---|---|---|---|
| Oliver et al. (2016) | USA (African American & Caribbean Black Adolescents) | 1,170 | National Survey of American Life (NSAL-Adolescent) | WHO-CIDI (Adolescent Version) | 9.2% | 7.0% | Adolescents aged 13-17; higher prevalence in Caribbean Black (12.4%) than African American (9%); strong comorbidity with anxiety; low treatment utilization |
| Silove et al. (2015) | Timor-Leste (Pregnant & Post-partum women) | 427 | Population-based study | Locally adapted DSM-IV explosive anger module | Not reported | 12% | Focused on explosive anger; high functional impairment; strongly associated with trauma, intimate partner violence, and adversity; not labeled as IED but comparable |
| Tay et al. (2022) | Myanmar Refugees in Malaysia | 2,058 | Population-based refugee survey | DSM-IV & DSM-5 CIDI Module | DSM-IV: N/A | DSM-IV: 5.9%, DSM-5: 3.4% | Three ethnic groups (Chin, Kachin, Rohingya); prevalence varied by group; high comorbidity with PTSD, depression, anxiety; higher prevalence than expected for refugee populations |
| Karam et al. (2008) | Lebanon (General population) | 2,857 | WMH Lebanon (LEBANON study) | WHO-CIDI 3.0 | 1.7% | Not reported | First national survey in Arab region; strong link between IED onset and war exposure (OR=12.72); high treatment gap; low prevalence compared to western countries. |

| | | | | | | | |
|---|---|---|---|---|---|---|---|
| Kessler et al. (2006) | USA (National Comorbidity Survey Replication - NCS-R) | 9,282 | National survey | WHO-CIDI 3.0 | 7.3% | 3.9% | One of the most cited prevalence studies globally; early onset (~14 years); high comorbidity with mood and substance disorders; low treatment rates for anger specifically. |
| Kim et al. (2010) | USA (Older Asian Americans) | 2,095 | NLAAS dataset | WHO-CIDI | Not reported | 2.3% | Focus on older (60+) Asian Americans; lower prevalence than younger Asians; explores age and cultural factors affecting prevalence; sample may have under-reporting due to social desirability. |
| Al-Hamzawi et al. (2012) | Iraq (National) | 4,332 | Iraq Mental Health Survey (IMHS) | WHO-CIDI | 1.7% | 1.5% | Strong exposure to war and violence; early onset (~18.5 years); high attack frequency (>140 lifetime attacks); high comorbidity with mood and anxiety but not substance disorders; remarkable persistence |
| Coccaro et al. (2005) | USA (Outpatient psychiatric sample) | 1,300 | Clinical outpatient sample | SCID-I/P | 6.3% | 3.1% | Early onset in adolescence, higher in males; comorbidity mainly with substance use disorder; 80% showed interest in treatment; first to highlight higher rates in clinical settings |

| | | | | | | | |
|---|---|---|---|---|---|---|---|
| Fincham et al. (2009) | South Africa (General population) | 4,351 | South African Stress and Health Study | WHO-CIDI | 9.5% (broad); 2.0% (narrow) | Not specified | First Sub-Saharan study; strong link to trauma exposure; higher prevalence in mixed-race and Caucasian participants; comorbid with mood, anxiety, and substance disorders; trauma dose-response observed |
| Gelegen & Tamam (2018) | Turkey (Psychiatric outpatients) | 406 | Outpatient clinical study | SCID + DSM-IV & DSM-5 | 16.7% (DSM-5) | 11.3% (DSM-5) | Higher prevalence in males (×3.8), rural residents, those with suicidal history; high comorbidity with ADHD, Conduct Disorder, ODD, and impulse control disorders. First Turkish IED study to use DSM-5. |
| McLaughlin et al. (2012) | USA (National adolescents) | 6,483 | NCS-A (National adolescent survey) | WHO-CIDI | 7.8% | 6.2% | Focused on adolescents (13–17 years); early onset (mean 12 years); high persistence (80% of lifetime cases were 12-month active); highly comorbid with mood, anxiety, and substance use disorders; low treatment rates specific for anger (6.5%). |
| Mundt et al. (2013) | Chile (Prison population) | 1,008 | National prison survey | CIDI 3.0 | Not reported | 5.7% | One of the few prison-focused studies; higher prevalence of IED in prisoners than general population; strong comorbidity with substance use, depression, and anxiety; confirms vulnerability of detained populations to IED. |

| | | | | | | | |
|---|---|---|---|---|---|---|---|
| Müller et al., 2011 | Germany (Psychiatric inpatients) | 234 | Clinical | SCID-ICD (DSM-IV) | 5.6% | 3.4% | European inpatient sample; focused on ICDs including IED; shows IED underdiagnosed by clinicians. |
| Tamam et al., 2011 | Turkey (Elderly outpatients) | 76 | Outpatient (Geriatric) | MIDI + DSM-IV | 15.8% | Not reported | First elderly-focused IED study; lifetime prevalence significantly higher in men (34.1% vs 8.6% in women). |
| Duan, 2010 | China (Shenzhen City) | 7,134 | Community | CIDI 3.1 (WHO-WMH) | 3.32% | 2.39% | Stratified random sample; early age of onset (~15 years); high comorbidity with MDD and OCD; low treatment rate (4.8%). |
| Rees et al., 2017 | Timor-Leste (Women in conflict areas) | 1,513 | Community | Adapted IED Module | Not reported | 12.2% | Conflict-affected women; strong associations with trauma, violence, and poverty; highly disabling. |
| Silove et al., 2018 | Timor-Leste (Perinatal women) | 427 | Community | Culturally adapted DSM-IV | Not reported | 12.0% | Pregnancy and postpartum women; related to IPV, trauma, and adversity; confirms Silove's earlier findings (2015). |
| Turner et al., 2021 | Germany (Youth offenders) | 161 | Prison (Youth) | CIDI & questionnaires | Not reported | 35.9% | Very high prevalence in young offenders; ACEs and externalizing disorders are key correlates. |

| | | | | | | | |
|---|---|---|---|---|---|---|---|
| Reardon et al., 2014 | USA (Veterans) | 232 | Clinical | DSM-IV (Structured Interview) | 24.0% | Not reported | High prevalence of IED in PTSD-affected veterans; IED linked with both externalizing and internalizing psychopathology. |
| Tay et al. (2015) | Papua New Guinea (West Papuan refugees) | 230 | Community-based refugee survey | Culturally adapted DSM-IV & DSM-5 modules | Not reported | 12% (LCA) | Latent class analysis identified an IED class comprising 12% of the sample. This group showed distinct symptoms of recurrent anger outbursts, aggression, and property destruction, without significant comorbidity with PTSD or depression. IED symptoms were strongly linked to long-term displacement and separation from family/homeland. The study supports the cross-cultural coherence of IED and highlights sociopolitical grievances as underlying causes. |
