## Appendix III for "Angry Without Borders: Global prevalence and factors of intermittent explosive disorder: A systematic review and meta-analysis"

| Study ID | Author(s), Year | Q1 | Q2 | Q3 | Q4 | Q5 | Q6 | Q7 | Q8 | Q9 | Total (0–9) |
|---|---|---|---|---|---|---|---|---|---|---|---|
| 1 | Pereira et al. (2020) | 1 | 1 | 1 | 1 | 1 | 0 | 1 | 1 | 1 | **8** |
| 2 | Coccaro et al. (2005) | 1 | 1 | 1 | 1 | 1 | 0 | 0 | 1 | 1 | **7** |
| 3 | Altwaijri et al. (2020) | 1 | 1 | 1 | 1 | 1 | 1 | 1 | 1 | 0 | **8** |
| 4 | Alhasnawi et al. (2009) | 1 | 1 | 1 | 0 | 1 | 0 | 0 | 1 | 0 | **5** |
| 5 | Ortega et al. (2008) | 1 | 1 | 1 | 1 | 1 | 0 | 1 | 1 | 1 | **8** |
| 6 | Shen et al. (2006) | 1 | 1 | 1 | 1 | 1 | 0 | 0 | 1 | 0 | **6** |
| 7 | Yoshimasu et al. (2011) | 1 | 1 | 1 | 1 | 1 | 0 | 1 | 1 | 1 | **8** |
| 8 | Scott et al. (2016) | 1 | 1 | 1 | 1 | 1 | 1 | 1 | 1 | 1 | **9** |
| 9 | Scott et al. (2020) | 1 | 1 | 1 | 1 | 1 | 1 | 1 | 1 | 1 | **9** |
| 10 | Oliver et al. (2016) | 1 | 1 | 0 | 1 | 1 | 0 | 1 | 1 | 0 | **6** |
| 11 | Silove et al. (2015) | 1 | 1 | 1 | 0 | 1 | 0 | 0 | 1 | 0 | **5** |
| 12 | Tay et al. (2022) | 1 | 1 | 1 | 1 | 1 | 1 | 1 | 1 | 1 | **9** |

| | | | | | | | | | | | |
|---|---|---|---|---|---|---|---|---|---|---|---|
| 13 | Karam et al. (2008) | 1 | 1 | 1 | 1 | 1 | 0 | 0 | 1 | 0 | **6** |
| 14 | Kessler et al. (2006) | 1 | 1 | 1 | 1 | 1 | 1 | 1 | 1 | 1 | **9** |
| 15 | Kim et al. (2010) | 1 | 1 | 1 | 1 | 1 | 0 | 0 | 1 | 0 | **6** |
| 16 | Al-Hamzawi et al. (2012) | 1 | 1 | 1 | 1 | 1 | 0 | 1 | 1 | 0 | **7** |
| 17 | Mundt et al. (2013) | 1 | 1 | 1 | 1 | 1 | 0 | 0 | 1 | 0 | **6** |
| 18 | Müller et al. (2011) | 1 | 1 | 0 | 1 | 1 | 0 | 0 | 1 | 0 | **5** |
| 19 | Tamam et al. (2011) | 1 | 1 | 0 | 1 | 1 | 0 | 0 | 1 | 0 | **5** |
| 20 | Duan (2010) | 1 | 1 | 1 | 1 | 1 | 0 | 1 | 1 | 0 | **7** |
| 21 | Rees et al. (2017) | 1 | 1 | 1 | 0 | 1 | 0 | 0 | 1 | 0 | **5** |
| 22 | Silove et al. (2018) | 1 | 1 | 1 | 0 | 1 | 0 | 0 | 1 | 0 | **5** |
| 23 | Turner et al. (2021) | 1 | 1 | 1 | 1 | 1 | 1 | 1 | 1 | 1 | **9** |
| 24 | Reardon et al. (2014) | 1 | 1 | 1 | 1 | 1 | 0 | 0 | 1 | 0 | **6** |

| | | | | | | | | | | |
|---|---|---|---|---|---|---|---|---|---|---|
| 25 | Tay et al. (2015) | 1 | 1 | 1 | 0 | 1 | 0 | 0 | 1 | 0 | **5** |
| 26 | Gelegen & Tamam (2018) | 1 | 1 | 1 | 1 | 1 | 0 | 1 | 1 | 1 | **8** |
| 27 | McLaughlin et al. (2012) | 1 | 1 | 1 | 1 | 1 | 1 | 1 | 1 | 1 | **9** |
| 28 | Fincham et al. (2009) | 1 | 1 | 1 | 1 | 1 | 0 | 0 | 1 | 0 | **6** |
| 29 | Coccaro et al. (2005) | 1 | 1 | 1 | 1 | 1 | 1 | 1 | 1 | 1 | **9** |

**High-quality studies (8–9 points)**: 12 (41.4%)

**Moderate quality (6–7 points)**: 11 studies (37.9%)

**Low quality (≤5 points)**: 6 studies (20.7%)
