## Appendix IV and Appendix V for "Angry Without Borders: Global prevalence and factors of intermittent explosive disorder: A systematic review and meta-analysis": Appendix IV_ied-funnel-plots-html.html

IED Prevalence Funnel Plots


### Publication Bias Assessment for IED Prevalence

Lifetime Prevalence Publication Bias

Egger's test: intercept = 8.45, p = .608  
Begg's test: τ = 0.74, p < .001

12-Month Prevalence Publication Bias

Egger's test: intercept = 7.91, p = .942  
Begg's test: τ = 0.80, p < .001

##### Interpretation:

These funnel plots assess publication bias in studies of Intermittent Explosive Disorder (IED) prevalence.
The vertical dotted lines represent pooled prevalence estimates (5.1% for lifetime and 4.4% for 12-month).
In an unbiased literature, points would be symmetrically distributed around the pooled estimate line,
forming an inverted funnel shape. Asymmetry suggests publication bias.

Both plots show significant Begg's test results (p < .001) indicating asymmetry,
while Egger's test results were non-significant. This suggests potential publication bias in IED prevalence literature.
